# T-cell activation, senescence, and exhaustion in asymptomatic HIV/Leish*mania infantum* co-infection

**DOI:** 10.1101/2023.03.06.23286828

**Authors:** Carolina de Oliveira Mendes-Aguiar, Manoella do Monte Alves, Amanda de Albuquerque Lopes Machado, Glória Regina de Góis Monteiro, Iara Marques Medeiros, Jose Wilton Queiroz, Iraci Duarte Lima, Richard D. Pearson, Mary E. Wilson, Marshall J. Glesby, Eliana Lúcia Tomaz do Nascimento, Selma Maria Bezerra Jerônimo

## Abstract

**Background:** *Leishmania infantum* is an opportunistic parasitic infection. An immunocompromised state increases the risk of converting asymptomatic infection to symptomatic visceral leishmaniasis (VL), which has a ∼5% fatality rate even with treatment. HIV coinfection increases the risk of death from VL.

**Methods:** A cross-sectional study was performed between 2014 and 2016 to determine the prevalence of *L. infantum* infection in HIV positive subjects residing in the state of Rio Grande do Norte, Brazil (n=1,372) and of these a subgroup of subjects were followed longitudinally. Subsequent incident cases of VL were ascertained from a public health database through 2018. A subgroup (n=69) of the cross-sectional study subjects was chosen to assess immune status (T cell activation, senescence, exhaustion) and outcome. The data were compared between asymptomatic HIV+/*L. infantum+* (HIV/Leish), symptomatic visceral leishmaniasis (VL), recovered VL, DTH+ (Delayed-Type Hypersensitivity response – Leishmanin skin test), AIDS/VL, HIV+ only (HIV+), and Non-HIV/Non *L. infantum* infection (control subjects).

**Results:** The cross-sectional study showed 24.2% of HIV+ subjects had positive anti-IgG *Leishmania* antibodies. After 3 years, 2.4% (8 of 333) of these HIV/Leish coinfected subjects developed AIDS/VL, whereas 1.05% (11 of 1,039) of HIV subjects with negative leishmania serology developed AIDS/VL. Poor adherence to antiretroviral therapy (p=0.0008) or prior opportunistic infections (p=0.0007) was associated with development of AIDS/VL. CD4+ (p=0.29) and CD8+ (p=0.38) T cells counts or viral load (p=0.34) were similar between asymptomatic HIV/Leish and HIV subjects. However, activated CD8^+^CD38^+^HLA-DR^+^ T cells were higher in asymptomatic HIV/Leish than HIV group. Likewise, senescent (CD57^+^) or exhausted (PD1^+^) CD8^+^ T cells were higher in asymptomatic HIV/Leish than in AIDS/VL or HIV groups.

**Conclusion:** Although asymptomatic HIV/Leish subjects had normal and similar CD4+ and CD8+ T cells counts, their CD8^+^T cells had increased activation, senescence, and exhaustion, which could contribute to risk of developing VL.

**Author Summary:** The frequency of asymptomatic HIV/Leish*mania infantum* (HIV/Leish) infection and the immunological status of subjects with HIV+ residing in the state of Rio Grande do Norte, Brazil, between 2014 and 2016 were studied. A high frequency of asymptomatic HIV/Leish*mania infantum* infection (HIV subjects with positive anti-IgG Leishmania antibodies) was found. Asymptomatic HIV/Leish subjects had CD8 T cells with higher markers of activation, senescence and exhaustion than the other groups (HIV-alone, symptomatic VL, Recovered VL, DTH+, AIDS/VL and Controls subjects). Poor adherence to antiretroviral therapy or history of previous opportunistic infection was associated with AIDS/VL. Asymptomatic HIV/Leish had high relative risk of developing AIDS/VL. Thus, subjects with HIV residing in endemic areas for VL should be assessed for their *L. infantum* infection status and advised to closely adhere to ART.

## Introduction

Visceral leishmaniasis (VL) is a prevalent disease in many countries, with more than 90% of cases in India, Nepal, Bangladesh, Sudan and Brazil. *Leishmania infantum* is the main etiologic agent of VL in Latin America and Europe [1]. *L. infantum* infection outcomes vary from asymptomatic to severe disease, but asymptomatic infection occurs in most people. In endemic regions of Brazil, a trend to harbor asymptomatic infection rather than VL has increased in parallel with improvement and nutrition, overall vaccination rate for children including measles and sanitary conditions around the household [2–4].

The factors determining the conversion from asymptomatic infection to VL are still not totally understood. Recent studies indicate that high antibody titers are more often see in subjects with symptomatic VL [5,6]. Subsets of T cells from asymptomatic *L. infantum* infected subjects are capable of activation after *Leishmania* stimulation, detected by the marker CD69^+^. The majority of asymptomatic subjects mount Th1-type cellular immune responses with IL-2 and IFN-*γ* production that is long-lasting if there is no immunosuppression, possibly indicating protective immunity [7]. In contrast, symptomatic VL subjects have unique T cell responses characterized by exhaustion and activation profiles, with decreased IFN-*γ* production [8].

The resurgence of VL in Europe occurred concurrently with the HIV epidemic, first spread through sharing of needles, and but also through the expansion of canine VL in several countries including Portugal, Spain, France and Italy. In Brazil, HIV was initially an infection of urban settings and VL was a disease predominantly in rural areas. However, in the last 30 years VL has become frequent in peri-metropolitan areas of major cities in the Northeast and Southeast regions of the country, and HIV has spread to rural areas. The increased regional overlap of both infections has led to changes in the epidemiology of VL due to co-infection [1,4]. In Rio Grande do Norte, this resulted in an increase in AIDS/VL cases detected after 2010 [3,4].

A high risk of VL is observed among people living with HIV [9], and *L. infantum* is an opportunistic pathogen in this population [10]. The impact of VL in people who are immunocompromised by HIV can be devastating, since both diseases suppress the immune system [11]. Asymptomatic co-infection with HIV and *Leishmania* increases the risk of VL development [12], and in Brazil co-infected patients have had more VL relapses and higher rates of death than singly infected persons [1]. Relapses of VL have been associated with falling CD4^+^ T cell counts [9].

*Leishmania* infection increases HIV viral load in serum from co-infected people [13]. Consistently, macrophages co-infected with HIV *in vitro* have higher *L. infantum* loads and increased capacity to uptake the mammalian stage, the amastigote [14,15]. Chronic HIV infection is known to lead to CD4^+^ T cell depletion and CD8^+^ T cell activation [11]. Cellular immune responses are essential for control of *Leishmania* spp. infection, with contributions of CD4^+^ T cells to the outcome of active disease and CD8+ cells contributing to immune protection. It is possible that the low CD4 T cells characterizing HIV infection synergize with the suppressed hematopoietic cell functions due to *Leishmania* spp. infection [16]. Indeed, AIDS/VL co-infected subjects exhibit increased CD8^+^ T cell activation during active VL disease and after remission compared to singly infected persons [17] and the chronic T cell activation phenotype is positively correlated with T cell senescence, accelerating immune cell impairment [16].

Our overarching hypothesis is that people living with asymptomatic HIV-*L. infantum* co-infection have higher risk of developing AIDS/VL than individuals with *L. infantum* infection alone. The goals of the study were to study the immune T cell activation, senescence and exhaustion in co-infected subjects, comparing AIDS/VL and asymptomatic HIV/Leishmania subjects with recovered VL, DTH+ (Montenegro skin test), AIDS/VL, HIV, and control subjects.

## Methods

### Study Design and population

A cross-sectional study was performed between July 2014 and January 2016, in which 1,372 HIV individuals residing in Rio Grande do Norte State, Brazil, which is endemic for VL, were tested for *L. infantum* infection by SLA and rK39 antigens using ELISA. A retrospective analysis was performed with data on symptomatic VL cases obtained from the Notifiable Diseases Information System (SINAN). All AIDS/VL reported cases in SINAN database were cross referenced to 1,372 HIV cases evaluated in cross-sectional study. All cases observed between July 2014 and December 2018 were analyzed. This was followed by the study of the immunological status of 69 subjects infected with HIV and/or Leishmania. The description of groups in the immunologic study is found in Table 1. All subjects included in immunological study had blood samples collected for further serological evaluation and for immunophenotyping, and their clinical data were collected. The subjects were questioned about adherence to ART therapy and previous opportunistic infections.

**Table 1.** Subject groups undergoing immunologic testing

| Group | Group name | n | Definition |
| --- | --- | --- | --- |
| HIV only | **HIV** | 16 | HIV subjects on ART, with negative *L. infantum* serology and no symptoms of VL |
| Asymptomatic HIV+/*L. infantum*+ | **Asympt HIV/Leish** | 10 | HIV subjects on ART, positive *L. infantum* serology and no symptoms of VL |
| AIDS/VL | **AIDS/VL** | 14 | AIDS and symptomatic VL* |
| Symptomatic VL | **VL** | 8 | Symptomatic VL* and HIV negative |
| Recovered VL | **Recovered VL** | 6 | Immunocompetent subjects treated for VL who had clinical recovery |
| DTH+ | **DTH+** | 9 | Immunocompetent subjects with DTH skin test response to *Leishmania* antigens with no history of VL |
| Healthy Control | **CS** | 6 | Healthy controls with no history of *Leishmania* or HIV infections |

\*The VL status was confirmed by either parasitological findings in the bone marrow aspirate or by presence of anti-*Leishmania* antibodies and with clinical findings consistent with the disease.

### ELISA serologic test for asymptomatic *Leishmania* case detection

HIV cases were screened for the presence of anti-Leishmania IgG antibodies using two *Leishmania infantum* antigens. (1) A soluble lysate of *L. infantum* (SLA) was produced from an isolate from a patient with VL in Natal that was typed in a World Health Organization Reference Laboratory (Elisa Cupolillo, Fiocruz, Rio de Janeiro, Brazil). (2) rK39 antigen for ELISA was kindly provided by Steven G. Reed (Infectious Diseases Research Institute, Seattle, WA). Both ELISA protocols have been previously described [18,19]. The cutoff for a positive reaction was determined as the mean + three standard deviations of the absorbance of negative control sera (minus absorbance in blank wells). Each serum sample was analyzed in duplicate. The results are shown as relative OD (rOD), normalized to the cutoff for each. The rOD was determined as the (mean of sample duplicates) – (mean absorbance in blank wells) / Cut-off. The results were expressed by rOD median/min-max; p value.

### Delayed Type Hypersensitivity skin test (DTH) to leishmania antigens

The antigen used for assessment of the DTH skin test was kindly provided by the Centro de Produção e Pesquisa de Imunobiológico/PR (CPPI, Paraná, Brazil). It was prepared from *L. amazonensis*, and it is the only DTH preparation approved for clinical use in Brazil. The DTH was placed and read in accordance with the ballpoint pen technique [20]. Induration greater than 5 mm in diameter was considered positive [21]

### Lymphocyte isolation and flow cytometric detection of T cell subsets

Two milliliters of peripheral blood were collected and red blood lysis buffer was added. After red blood cell lysis, white cells were washed by centrifugation in 0.1% PBS-Azide and then labeled with monoclonal antibodies. Activation status (CD38-FITC; BD, clone HIT2/HLA-DR-PE; Ebioscience, clone L243), Senescence (CD57-FITC; BD, clone NK-1) and Exhaustion (PD-1-PERCP; Biolegend, clone NAT105) were analyzed in CD4^+^CD3^+^ and in CD8^+^CD3^+^ T cells by flow cytometry. After antibody incubation, cells were fixed in 1% formol buffer. Thirty thousand events were acquired from each sample using a FACS Canto II flow cytometer (BD, Canto II flow cytometer, Becton Dickinson Bioscience, USA). FlowJo vX software was used for flow cytometry analysis. Lymphocytes were gated in forward scatter and side scatter dot plot graphic. Activation, senescence and Exhaustion markers were evaluated in gated CD4^+^CD3^+^ and in CD8^+^CD3^+^ T lymphocyte subpopulation. Results were expressed as percentage (median; min-max).

### Ethical considerations

The protocol and informed consent were reviewed and approved by the Federal University of Rio Grande do Norte Ethical Committee (CEP-UFRN) and by the Brazilian National Ethics Committee (CONEP). The certificate of ethical approval is CAAE 12675013.7.0000.5537. Informed consent was obtained, and the consent form was signed by the participant or their legal guardian.

### Statistical analysis

Continuous variables are expressed as medians and interquartile ranges. The statistical analyses were performed using GraphPad Prism software (version 8.0, San Diego, CA, USA). Kruskal-Wallis nonparametric ANOVA with multiple comparisons was used to analyze differences between groups. Differences were considered statistically significant when the p value was <0.05. Chi-square test was performed to compare categorical variables. Relative risk was calculated using omni calculator (https://www.omnicalculator.com/statistics/relative-risk) with 95% confidence intervals (Cl).

## Results

### 1. Leishmania infection in people living with HIV in endemic area of Rio Grande do Norte, Brazil, and visceral leishmaniasis development

People living with HIV (n=1,372) were tested for *L. infantum* infection, using SLA and rK39 anti-IgG ELISA serology. Of those, 333 people with HIV (24.2%, 333/1,372) had positive antibodies to rK39 and/or SLA. High titers of SLA or rK39 were detected in 17.2% (236/1,372) or 12.9% (178/1,372) patients, respectively. SLA rOD results correlated with rK39 rDO (r=0.82; p<0.001). Three years after the initial screening, 19 (1.38%) subjects developed visceral leishmaniasis (AIDS/VL) and were treated for VL. Among the subjects who developed AIDS/VL, 11 had initially tested negative for Leishmania infection (11 of 1,039; 1.05%) and 8 had tested positive for Leishmania (8 of 333; 2.4%) (Chi-square; p=0.033) (Table 2).

**Table 2.** Relative risk of HIV subjects to development AIDS/VL

|  | HIV | HIV/SLA-* | HIV/SLA+*<br>(asymptomatic) | Relative<br>Risk | Chi-<br>square |
| --- | --- | --- | --- | --- | --- |
| AIDS/VL<br>outcome | 19/1,372<br>(1.38%) | 11/1,039<br>(1.05%) | 8/333<br>(2.4%) | 2.27 | **0.033** |

\*Relative risk – comparison between HIV/SLA- and HIV/SLA+ groups

### 2. Immunologic and virologic assessments comparing HIV and HIV-asymptomatic Leishmania infected group

The immunologic and virologic status of a subgroup of asymptomatic HIV/Leish subjects was compared to HIV, AIDS/VL, VL, Recovered VL, DTH+ and CS subjects. Results are shown in Table 3, as median (min-max); and in Tables 4 and 5 as *p values*. Most of the subjects were males as VL (87.5%), AIDS/VL (85%) and asymptomatic HIV/Leish individuals (70%) (Table 3). There was no difference in the age between groups (Kruskal-Wallis p=0.56) (Table 3).

**Table 3.** Clinical and laboratory characteristics of patients included in the immunologic study

| Groups | Age<br>(Years) | Gender<br>(%male) | CD4<sup>+</sup> T cells<br>count<br>(cells/mm<sup>3</sup>) | CD8<sup>+</sup> T cells<br>count<br>(cells/mm<sup>3</sup>) | HIV Viral load<br>(Log<sub>10</sub>copies/mL) | rOD anti-<br>IgG SLA | rOD anti-<br>IgG rK39 |
| --- | --- | --- | --- | --- | --- | --- | --- |
| CS<br>(n=6) | 45<br>(35-52) | 3F/3M<br>(50%) | ND | ND | ND | 0.15<br>(0.13-0.7) | 0.29<br>(0.15-0.7) |
| HIV<br>(n=16) | 42<br>(21-54) | 7M/9F<br>(43,7%) | 620<br>(64-1,153) | 817<br>(284-1,992) | UND<br>(UND-3.9) | 0.17<br>(0.14-0.49) | 0.33<br>(0.16-0.61) |
| Asympt<br>HIV/Leish<br>(n=10) | 41<br>(29-62) | 7M/3F<br>(70%) | 414<br>(105-747) | 1002<br>(316-35,462) | UND<br>(UND-4.5) | 1.66<br>(1.01-3.86) | 0.96<br>(0.35-2.97) |
| AIDS/VL<br>(n=14) | 40<br>(18-53) | 12M/2F<br>(85%) | 85<br>(8-161) | 737<br>(218-1,194) | 5.3<br>(UND-7.0) | 0.55<br>(0.13-6.17) | 0.59<br>(0.19-7.91) |
| VL<br>(n=8) | 35<br>(16-51) | 7M/1F<br>(87,5%) | ND | ND | ND | 3.93<br>(1.63-10.47) | 5.79<br>(1.02-9.47) |
| Recovered<br>VL (n=9) | 28<br>(15-32) | 2F/4M<br>(66.67%) | ND | ND | ND | 1.35<br>(0.44-3.5) | 1.5<br>(0.57-14.88) |
| DTH+<br>(n=6) | 36<br>(15-56) | 4M/5F<br>(44.5%) | ND | ND | ND | 1.03<br>(0.66-1.22) | 1.7<br>(1.17-3.6) |

Results are expressed in Median (min-max); F – female; M – Male; ND – not done; UND – undetectable (<40 viral copies/mL); rOD – relative OD.

**Table 4.**
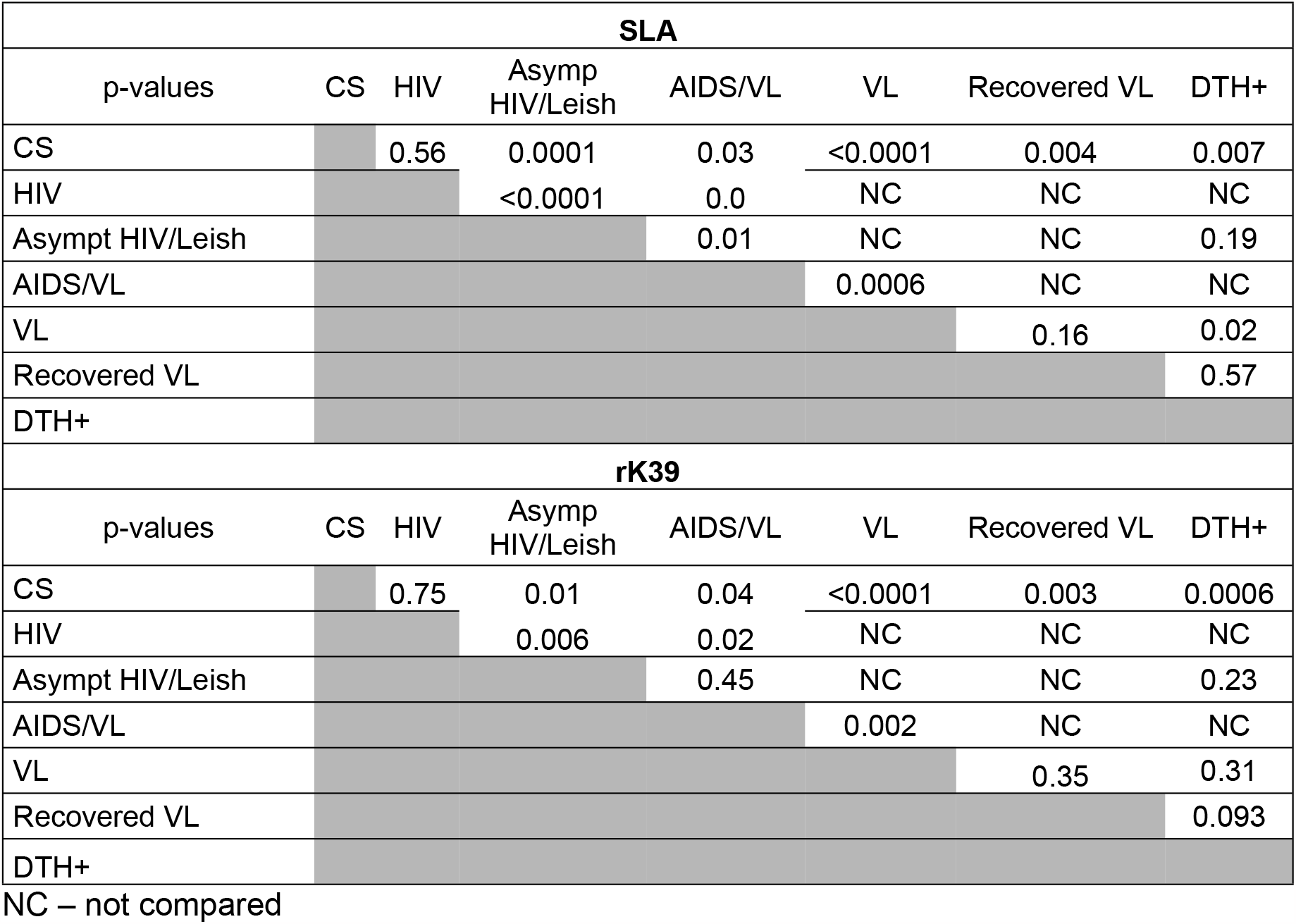
Comparison of anti-Leishmania antibodies among the groups with respective *p values*

| SLA |  |  |  |  |  |  |  |
| --- | --- | --- | --- | --- | --- | --- | --- |
| p-values | CS | HIV | Asymp HIV/Leish | AIDS/VL | VL | Recovered VL | DTH+ |
| CS |  | 0.56 | 0.0001 | 0.03 | <0.0001 | 0.004 | 0.007 |
| HIV |  |  | <0.0001 | 0.0 | NC | NC | NC |
| Asympt HIV/Leish |  |  |  | 0.01 | NC | NC | 0.19 |
| AIDS/VL |  |  |  |  | 0.0006 | NC | NC |
| VL |  |  |  |  |  | 0.16 | 0.02 |
| Recovered VL |  |  |  |  |  |  | 0.57 |
| DTH+ |  |  |  |  |  |  |  |
| rK39 |  |  |  |  |  |  |  |
| p-values | CS | HIV | Asymp HIV/Leish | AIDS/VL | VL | Recovered VL | DTH+ |
| CS |  | 0.75 | 0.01 | 0.04 | <0.0001 | 0.003 | 0.0006 |
| HIV |  |  | 0.006 | 0.02 | NC | NC | NC |
| Asympt HIV/Leish |  |  |  | 0.45 | NC | NC | 0.23 |
| AIDS/VL |  |  |  |  | 0.002 | NC | NC |
| VL |  |  |  |  |  | 0.35 | 0.31 |
| Recovered VL |  |  |  |  |  |  | 0.093 |
| DTH+ |  |  |  |  |  |  |  |

NC – not compared

**Table 5.** Comparison of HIV load and CD4 counts

| Viral load |  |  |  |
| --- | --- | --- | --- |
| p-values | HIV | Asymp HIV/Leish | AIDS/VL |
| HIV |  | 0.34 | <0.0001 |
| Asympt HIV/Leish |  |  | 0.004 |
| AIDS/VL |  |  |  |
| CD4 counts |  |  |  |

| p-values | HIV | Asympt<br>HIV/Leish | AIDS/VL |
| --- | --- | --- | --- |
| HIV |  | 0.29 | <0.0001 |
| Asympt HIV/Leish |  |  | 0.003 |
| AIDS/VL |  |  |  |

NC – not compared

All asympt HIV/Leish individuals (10 of 10) had positive anti-IgG SLA serology, and 5 of 10 individual had positive anti-IgG rK39 serology. Asympt HIV/Leish individuals had higher anti-SLA IgG antibodies and similar levels of anti-rK39 IgG antibodies compared to AIDS/VL (Table 3 and 4). VL patients had higher levels of anti-Leishmania antibodies than AIDS/VL group (SLA and rK39) or DTH+ subjects (SLA). Most AIDS/VL subjects had negative serology to *Leishmania infantum* antigens (SLA: 71.4%/ rK39: 78.5%). DTH+ subjects had similar rOD antibodies levels as asympt HIV/Leish individuals or recovered VL subjects (Table 3 and 4).

HIV groups differed with respect to their HIV viral loads and CD4+ T cell counts, but not their CD8+ T cell counts (Table 3 and 5). HIV and asympt HIV/Leish individuals had lower viral loads than individuals with AIDS/VL (Table 3 and 5). At the same time, CD4+ T counts were higher in HIV and in asympt HIV/Leish individuals than in AIDS/VL (table 3 and 5). Viral load, and CD4 and CD8 T cell counts did not differ between HIV and asympt HIV/Leish individuals (Table 3 and 5). HIV viral load correlated negatively to CD4 T cell counts (r=-0.53; p=0.005). No differences in CD8+ T cell counts were observed between the HIV, asympt HIV/Leish and AIDS/VL groups (ANOVA Kruskal-Wallis p=0.25) (table 3).

ART was used by 81.25% (13/16) of the HIV subjects (Table 6). However, in *Leishmania* infected groups, the adherence to ART was lower. Sixty percent (6/10) of asympt HIV/Leish subjects and 7.15% (1/14) of AIDS/VL subjects used ART regularly (Table 6). Subjects were asked about their history of prior chronic infections with *Toxoplasma* spp., *Mycobacterium tuberculosis* or *Mycobacterium leprae*. Before enrollment, 25% (4/16) of HIV subjects had had these chronic infections, 40% (4/10) of asympt HIV/Leish subjects had had previous chronic infections, and 84% (11/14) of AIDS/VL cases had had previous infections (Table 6). Chi-square analysis revealed that both the incorrect use of ART therapy (Fisher Chi-square 0.0008) and a history of previous chronic infection (Fisher Chi-square 0.0007) were associated with an increased risk of AIDS/VL outcome (Table 6).

**Table 6.**
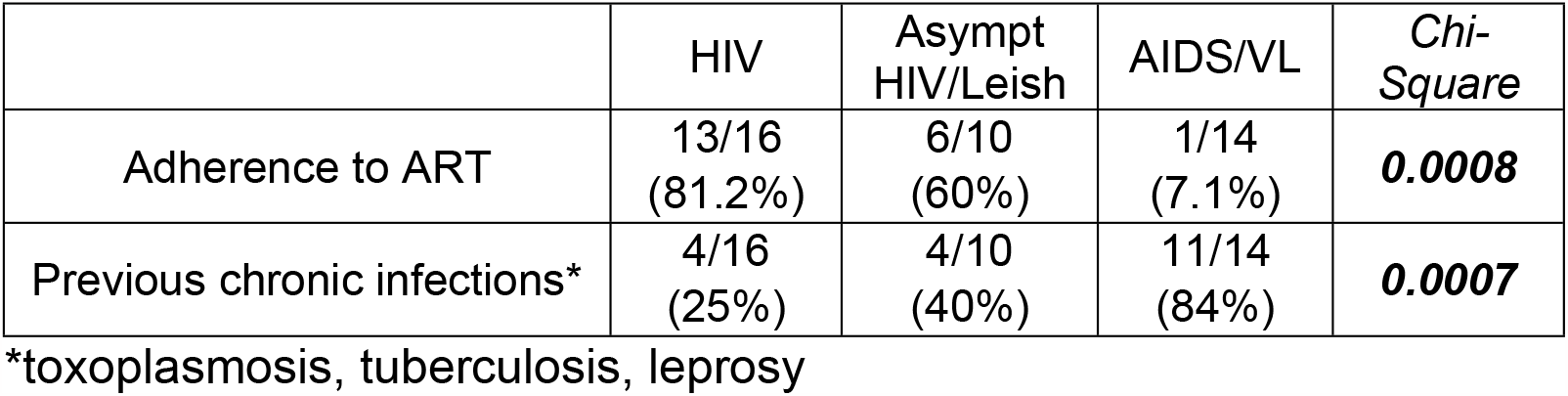
Adherence to ART treatment or previous infection and AIDS/VL outcome association

|  | HIV | Asympt<br>HIV/Leish | AIDS/VL | Chi-Square |
| --- | --- | --- | --- | --- |
| Adherence to ART | 13/16<br>(81.2%) | 6/10<br>(60%) | 1/14<br>(7.1%) | **0.0008** |
| Previous chronic infections* | 4/16<br>(25%) | 4/10<br>(40%) | 11/14<br>(84%) | **0.0007** |

\*toxoplasmosis, tuberculosis, leprosy

### 3. Asymptomatic HIV/Leish*mania infantum* infection increased activation, senescence and exhaustion immunological status in CD8^+^T cells

T cell activation, senescence and exhaustion were examined in HIV-subjects. The results are shown in table 7 and 8, and Figures 1 and 2.

**Table 7.** Proportions of subjects with makers of immune activation, senescence or exhaustion in T cell subsets

|  | CD3<sup>+</sup>CD4<sup>+</sup> T cell |  |  | CD3<sup>+</sup>CD8<sup>+</sup> T cell |  |  |
| --- | --- | --- | --- | --- | --- | --- |
|  | CD38<sup>+</sup>HLA-DR<sup>+</sup> | CD57<sup>+</sup> | PD1<sup>+</sup> | CD38<sup>+</sup>HLA-DR<sup>+</sup> | CD57<sup>+</sup> | PD1<sup>+</sup> |
| CS<br>(n=6) | 2.0%<br>(0.8-13.7) | 12.87%<br>(1.27-37.0) | 8.65%<br>(3.9-12.9) | 2.42%<br>(0.54-3.58) | 39.4%<br>(15.9-60.1) | 0.04%<br>(0.0-0.55) |
| HIV<br>(n=16) | 2.52%<br>(0.78-17.7) | 4.1%<br>(0.73-52.2) | 5.1%<br>(0.18-15.4) | 4.19%<br>(2.14-25.1) | 39.1%<br>(17.1-69.6) | 2.52%<br>(0.01-58.0) |
| Asympt<br>HIV/Leish<br>(n=10) | 6.89%<br>(3.31-15.8) | 15.65%<br>(2.6-66.7) | 5.37%<br>(1.3-21.4) | 24.55%<br>(6.65-50.4) | 62.8%<br>(42.6-88.0) | 42.6%<br>(15.7-80.1) |
| AIDS/VL<br>(n=14) | 18.89%<br>(0.74-86.2) | 10.88%<br>(0.2-40.0) | 18.0<br>(0.4-40.2) | 36.85%<br>(14.7-96.9) | 54.6%<br>(21.3-72.4) | 21.45%<br>(6.2-57.0) |
| VL<br>(n=8) | 14.2%<br>(0.85-31.7) | 5.81%<br>(1.2-28.7) | 11.7%<br>(0.7-45.9) | 54.05%<br>(2.08-65.0) | 49.7%<br>(11.1-59.1) | 15.15%<br>(4.32-55.7) |
| Recovered<br>VL (n=9) | 2.7%<br>(1.34-7.1) | 6.82%<br>(3.0-47.9) | 18.95%<br>(5.93-40.1) | 5.41%<br>(2.77-12.5) | 43.6%<br>(11.2-71.2) | 1.55%<br>(0.28-12.2) |
| DTH+<br>(n=6) | 0.39%<br>(0.02-1.83) | 8.3%<br>(0.71-17.6) | 4.02%<br>(1.82-30.9) | 0.08%<br>(0.0-7.42) | 35.1%<br>(12.0-59.6) | 0.24%<br>(0.0-1.69) |

Results were expressed Median (Min-Max)

**Table 8.** Comparison of T cell markers among groups

| CD4<sup>+</sup> T cell activation markers (HLA-DR, CD38) |  |  |  |  |  |  |  |
| --- | --- | --- | --- | --- | --- | --- | --- |
| p-values | CS | HIV | Asympt<br>HIV/Leish | AIDS/VL | VL | Recovered<br>VL | DTH+ |
| CS |  | 0.99 | 0.11 | **0.0008** | 0.05 | 0.96 | 0.09 |
| HIV |  |  | **0.04** | **0.0004** | NC | NC | NC |
| Asympt HIV/Leish |  |  |  | 0.25 | NC | NC | **0.0002** |
| AIDS/VL |  |  |  |  | 0.57 | NC | NC |
| VL |  |  |  |  |  | 0.06 | **<0.0001** |
| Recovered VL |  |  |  |  |  |  | 0.08 |
| DTH+ |  |  |  |  |  |  |  |
| CD8<sup>+</sup> T cell activation markers (HLA-DR, CD38) |  |  |  |  |  |  |  |
| p-values | CS | HIV | Asympt<br>HIV/Leish | AIDS/VL | VL | Recovered<br>VL | DTH+ |
| CS |  | 0.11 | **0.0007** | **<0.0001** | **0.0001** | 0.95 | 0.18 |
| HIV |  |  | **0.01** | **0.0001** | NC | NC | NC |
| Asympt HIV/Leish |  |  |  | 0.35 | NC | NC | **0.0001** |
| AIDS/VL |  |  |  |  | 0.84 | NC | NC |
| VL |  |  |  |  |  | **0.01** | **<0.0001** |
| Recovered VL |  |  |  |  |  |  | 0.13 |
| DTH+ |  |  |  |  |  |  |  |
| **CD4+ T cell senescence marker (CD57)** |  |  |  |  |  |  |  |
| p-values | CS | HIV | Asympt HIV/Leish | AIDS/VL | VL | Recovered VL | DTH+ |
| CS |  | 0.28 | 0.55 | 0.71 | 0.45 | 0.95 | 0.38 |
| HIV |  |  | **0.04** | **0.05** | NC | NC | NC |
| Asympt HIV/Leish |  |  |  | 0.74 | NC | NC | 0.09 |
| AIDS/VL |  |  |  |  | 0.19 | NC | NC |
| VL |  |  |  |  |  | 0.49 | 0.91 |
| Recovered VL |  |  |  |  |  |  | 0.41 |
| DTH+ |  |  |  |  |  |  |  |
| **CD8+ T cell senescence marker (CD57)** |  |  |  |  |  |  |  |
| p-values | CS | HIV | Asympt HIV/Leish | AIDS/VL | VL | Recovered VL | DTH+ |
| CS |  | 0.98 | 0.01 | 0.23 | 0.57 | 0.57 | 0.88 |
| HIV |  |  | 0.002 | 0.11 | NC | NC | NC |
| Asympt HIV/Leish |  |  |  | 0.12 | NC | NC | 0.004 |
| AIDS/VL |  |  |  |  | 0.54 | NC | NC |
| VL |  |  |  |  |  | 0.96 | 0.42 |
| Recovered VL |  |  |  |  |  |  | 0.43 |
| DTH+ |  |  |  |  |  |  |  |
| **CD4+ T cell exhaustion marker (PD1)** |  |  |  |  |  |  |  |
| p-values | CS | HIV | Asympt HIV/Leish | AIDS/VL | VL | Recovered VL | DTH+ |
| CS |  | 0.31 | 0.91 | 0.19 | 0.41 | 0.08 | 0.86 |
| HIV |  |  | 0.18 | **0.002** | NC | NC | NC |
| Asympt HIV/Leish |  |  |  | 0.16 | NC | NC | 0.75 |
| AIDS/VL |  |  |  |  | 0.65 | NC | NC |
| VL |  |  |  |  |  | 0.31 | 0.27 |
| Recovered VL |  |  |  |  |  |  | 0.04 |
| DTH+ |  |  |  |  |  |  |  |
| **CD8+ T cell exhaustion marker (PD1)** |  |  |  |  |  |  |  |
| p-values | CS | HIV | Asympt HIV/Leish | AIDS/VL | VL | Recovered VL | DTH+ |
| CS |  | **0.05** | **<0.0001** | **0.0001** | **0.002** | 0.16 | 0.54 |
| HIV |  |  | **0.003** | **0.01** | NC | NC | NC |
| Asympt HIV/Leish |  |  |  | 0.19 | NC | NC | **<0.0001** |
| AIDS/VL |  |  |  |  | 0.61 | NC | NC |
| VL |  |  |  |  |  | 0.12 | **0.006** |
| Recovered VL |  |  |  |  |  |  | 0.35 |
| DTH+ |  |  |  |  |  |  |  |

NC – not compared

**Figure 1:**
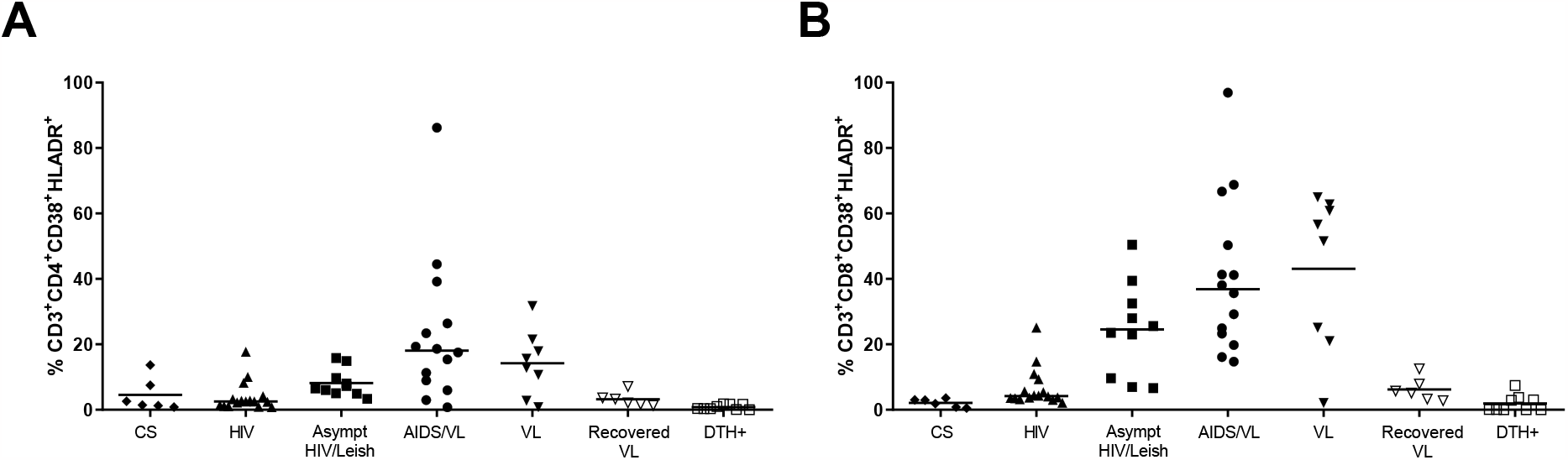
Immune activation CD38^+^ HLA-DR^+^ status in CD3^+^CD4^+^ (A) and in CD3^+^CD8^+^ T cells (B). Each point in the graphs represents one individual. Significance: p<0.05.

**Figure 2.**
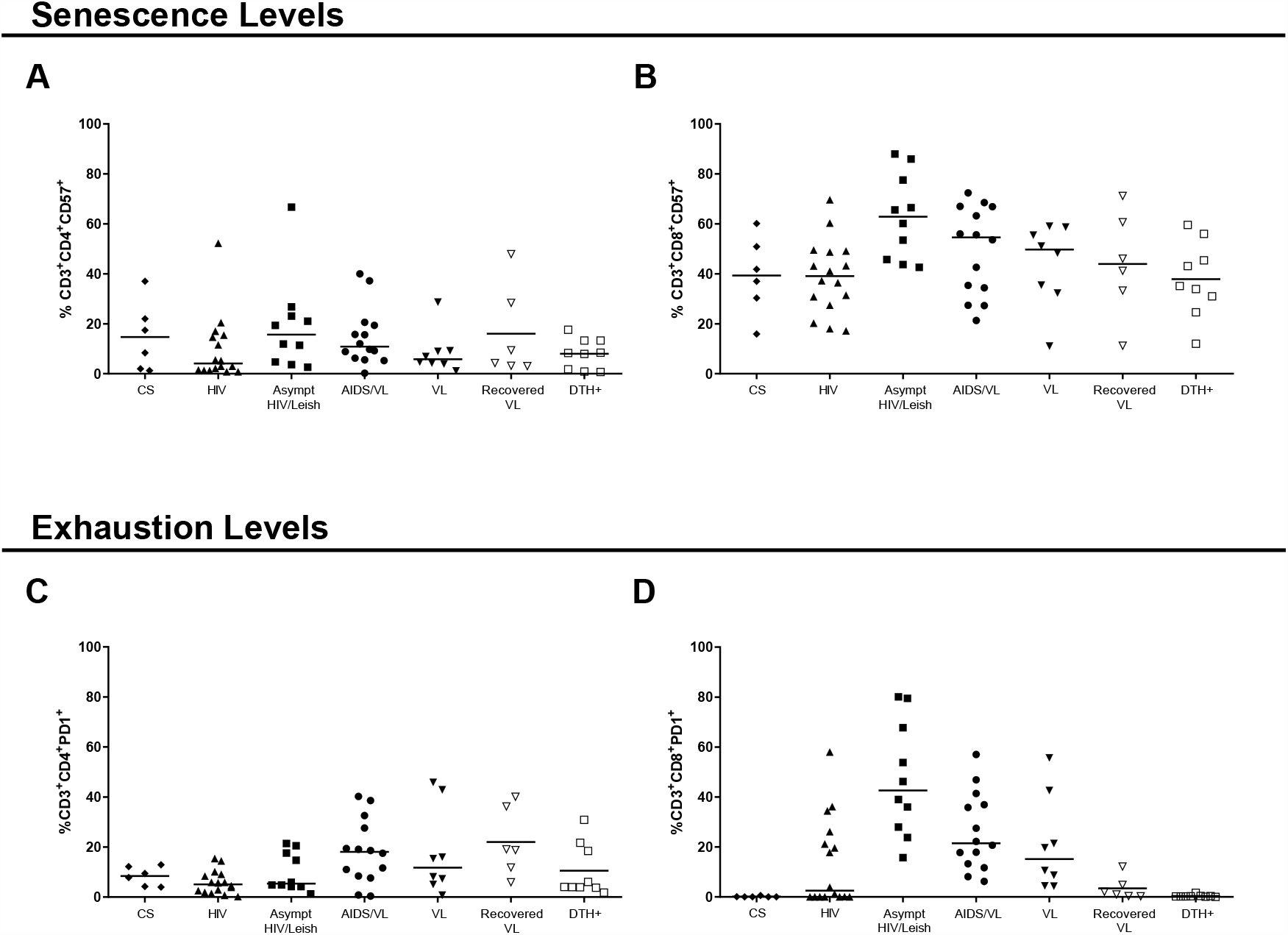
Proportions of cells positive for markers of senescence (CD57^+^) and exhaustion (PD1^+^) in CD3^+^CD4^+^ (A and C) or in CD3^+^CD8^+^ T cells (B and D). Each point in the graphs represents one individual. Significance: p>0.05.

CD38 is a marker of cell activation. In HIV infection, it predicts HIV progression and correlates with high levels of viral load [1]. High levels of activated T cells were observed in VL, AIDS/VL and asympt HIV/Leish subjects, using CD38^+^ and HLA-DR^+^ expression as markers of activation *ex-vivo* (Fig 1 and Table 7 and 8). VL subjects had higher proportions of activated T-cells than DTH+. After cure, recovered VL subjects had comparatively lowered proportions of CD8^+^CD38^+^HLA-DR^+^ activated T cells than those with VL (Fig 1 and Table 7 and 8). Similar higher and activation levels were observed between VL and AIDS/VL subjects, in both CD4^+^ and CD8^+^ T cells. VL patients had higher proportions of CD38+HLA-DR+ T cells than CS. There were no differences between T-cell activation in CS subjects compared to HIV subjects, recovered VL or DTH+ subjects. AIDS/VL had higher proportions of activated T cells than HIV or CS (Fig 1; Tables 7 and 8). Asympt HIV/Leish subjects had higher proportions of activated CD8+ T cells than CS subjects (Fig 1; Tables 7 and 8); and also showed higher proportions of activated CD4^+^ T cells than HIV or DTH+. Proportions of T cells expressing activation markers did not differ asympt HIV/Leish and AIDS/VL subjects in CD4^+^ or CD8^+^ T cells. However, proportions of CD8^+^ T cells expressing activation markers were higher for asympt HIV/Leish subjects compared to HIV or DTH+ (Fig 1; Tables 7 and 8). CD8^+^CD38^+^ T cells seem to play a role in VL and HIV, especially in AIDS/VL and asymptomatic HIV/Leishmania co-infected subjects.

CD57 is one of the senescence associated molecules. Senescent cells have shorter-ending telomers, that affect cellular function and proliferation. Cancer, chronic infections, autoimmune disease and aging exhibit high levels of senescent cells [22]. Similar percentages of senescent CD4^+^T cells expressing CD57^+^ were observed between most analyzed groups (Fig 2A; Tables 7 and 8). This includes a lack of significant difference between senescence in CD4^+^ and CD8^+^ T cells of asympt HIV/Leish and AIDS/VL subjects. Nonetheless there were significantly lower proportions of senescent CD4^+^T cells in HIV group than asympt HIV/Leish or in AIDS/VL subjects (Fig 2A; Tables 7 and 8). Proportions of senescent CD8^+^T cells were significantly different across the HIV and VL groups (Fig 2A; Tables 7 and 8). Asympt HIV/Leish expressed higher proportions of CD57^+^CD8+ T cells than CS, HIV and DTH+ groups. Similar levels of senescent TCD8+ cells were observed in subjects with AIDS/VL, HIV, asympt, HIV/Leish or VL. Even after cure, recovered VL subjects showed similar levels of senescent CD8 T cells compared to VL patients (Fig 2A; Tables 7 and 8). Thus, more CD4^+^ and CD8^+^ T cells became senescent in persons with Leishmania and HIV co-infection than in asymptomatic HIV/Leish subjects.

Chronic T activation, such as occurs in HIV infection or visceral leishmaniasis, leads to cell exhaustion. Exhausted cells are dysfunctional and have reduced responses to antigen. PD-1 (CD279) is mainly associated exhaustion [22]. Proportions of CD4^+^T expressing this marker of exhaustion in AIDS/VL subjects were significantly higher than among HIV subjects (Fig 2B; Tables 7 and 8). Remarkably there were few other significant differences in CD4+ T cell exhaustion markers between the groups (Fig 2B; Tables 7 and 8). In contrast to CD4+ T cells, the overall proportions of CD8^+^T cells expressing the exhaustion marker differed significantly across the groups (Fig 2B; Tables 7 and 8). Proportions were highest in the asympt HIV/Leish group, which differed from proportions in HIV, DTH+ or CS subjects. Levels of CD8+ T cell exhaustion were also high in HIV, AIDS/VL and VL subjects. AIDS/VL had similar proportions of exhausted CD8^+^ T compared to VL or asympt HIV/Leish subjects (Fig 2B; Tables 7 and 8). However, AIDS/VL had higher proportions than HIV or CS. VL patients had higher senescent CD8^+^T cells than DTH+ or CS. After cure of VL (recovered VL), the levels of CD8^+^PD1^+^ T cells were similar to those in CS and DTH+ subjects (Fig 2B; Tables 7 and 8). CS subjects had lower percentages of senencent CD8^+^T cells than HIV subjects. Thus, CD8^+^T cells of HIV/*L. infantum* subjects seem to be compromised with high senescence and exhaustion levels.

## Discussion

The urbanization of *L. infantum* in northeastern Brazil and the spread of HIV have contributed to the likelihood of concurrent infections. Since 2010, an increase in AIDS/VL cases as well as in asymptomatic *L. infantum* infection have been observed in the outskirts of Natal, as shown in this study, and in other Brazilian cities [23,24].

Herein we report a high prevalence (24.2%) of *L. infantum* positive serology among HIV-infected residents of Natal. Asymptomatic HIV/Leishmania prevalence reported from other endemic areas for VL has ranged from 3% to 25% [25–27]. Differences in the sensitivity and specificity of *Leishmania* detection tests can affect the calculated prevalence of asymptomatic HIV/*Leishmania* co-infection [28]. Sensitive PCR showed high prevalence of asymptomatic HIV/*Leishmania* co-infection in Italy [25]. In our study, rK39 and SLA ELISA were used to screen the HIV+ population. The results in this study are similar to those previously observed from Natal [19]. It is well known that AIDS/VL patients have lower titers of anti-Leishmania antibodies than persons with VL alone [29]. Of note, asymptomatic HIV/Leish (HIV subject with no clinical or laboratorial sign of *L. infantum* infection but positive serology) have an increased risk of developing AIDS/VL.

Asymptomatic HIV/Leish and AIDS/VL subjects in this study were predominately males. The prevalence of HIV is higher among males than females in the state of Grande do Norte. But VL has also been recognized more often in males [4,30]; while both sexes have been shown to be equally exposed to *L. infantum* infection in the region [31]. Sexual hormones seem to play a role in the pathogenesis and progression to VL, once males reach puberty. And higher levels of IL10 and TNF have been observed in male than female mice infected with *Leishmania infantum* [32].

The consequence of HIV and *Leishmania infantum* co-infection is an increased likelihood of progression to AIDS/VL,[1]. The negative correlation between CD4 T cells counts and HIV viral load suggests a spectrum among HIV-infected subjects from asymptomatic HIV/Leish to AIDS/VL. In our study persons with asymptomatic HIV/Leish had similar CD4 and CD8 T cells counts and HIV viral loads as HIV subjects. The lack of adherence to ART therapy or a history of prior infectious diseases were associated with an increased risk of progressing to AIDS/VL. Adherence to ART could be maintaining protective Th1 cytokine production by CD4^+^T cells [33].

T cell activation is strongly associated with progression to AIDS and death [34]. In the present study chronic immune activation was observed in AIDS/VL subjects in comparison to HIV+ or negative subjects. The positive correlation between *Leishmania* infection and higher levels of CD8^+^T activation in HIV-co-infected subjects in comparison to those with HIV alone suggests that *L. infantum* plays a role in CD8+T cell activation during co-infections [35].

*Leishmania infantum* infection causes several immunological alterations contributing to immune suppression. Both VL and AIDS/VL subjects had higher percentages of *ex vivo* activation. Persons with symptomatic leishmaniasis have previously been shown to have higher percentages of *ex vivo* activation, but it is not *Leishmania* specific, since CD4^+^T and CD8^+^T cells exposed to leishmanial antigen did not express activation molecules [8]. Despite lower *ex vivo* activation observed in DTH+ or recovered-VL subjects, these groups mounted specific T cell response after stimulation [8]. It is probably that asymptomatic HIV/Leishmania subjects can mount a specific Th1 response to SLA antigen [36], since asymptomatic HIV/Leish subjects showed similar percentage of activated CD4^+^T when compared to HIV or DTH+ individuals, and lower CD4^+^T levels of activation than AIDS/VL subjects.

Higher percentages of senescent CD8^+^T(CD3^+^CD8^+^CD57^+^) or exhausted (PD1^+^) CD4^+^ or CD8^+^ T cells were found in AIDS/VL than in HIV or control subjects, illustrating the deleterious impact of dual HIV and *Leishmania* infections, increasing senescence and exhaustion, and consequently, the deterioration of the immune system [1]. In this study, higher percentages of senescence (CD57^+^) and exhaustion (PD1^+^) were observed in the VL and AIDS/VL groups. But senescence was not affected in immunocompetent groups, suggesting that it was caused by HIV. During active VL, more exhausted CD8^+^T cells were observed than in persons who had recovered from VL or DTH+ subjects. Exhausted CD8^+^T cells from active VL patients did not produce IFN-*γ* in response to specific stimulation; however, this suppression was transitory, since after treatment, CD8^+^T cells recover the capacity of specific response [37].

Asymptomatic HIV/Leish co-infected subjects seemed to be controlling both infections. This group did not have any VL symptoms and had similar HIV viral load, CD4 and CD8 T cell counts in comparison to HIV subjects. These similarities in clinical and laboratory findings of asymptomatic HIV/Leish co-infection may be due to properly CD4^+^T cells counts and HIV viral load [25]. But chronic active *Leishmania* infection can stimulate immune activation [17] leading to increase HIV viral load and accelerating progression to AIDS/VL [11,38].

The management of asymptomatic HIV/Leishmania co-infected subjects starts with optimizing ART therapy and close follow-up of clinical status, viral load and CD4 count. Persons who develop symptoms of VL should be treated with liposomal amphotericin or alternative anti-leishmanial therapy. The role of anti-leishmanial therapy in the asymptomatic co-infected person is uncertain. Further studies are needed to determine its role, particularly in persons with low or falling CD4 counts or uncontrolled viral loads.

## Data Availability

All relevant data are within the manuscript and its Supporting Information files.

## Acknowledegment

We are thankful to the health personnel from Giselda Trigueiro Hospital, Central Laboratory (Laboratório Central de Saúde Pública do Rio Grande do Norte Dr. Almino Fernandes) for helping with the volunteers’ recruitment.

